# Auditor Models to Suppress Poor AI Predictions Can Improve Human-AI Collaborative Performance

**DOI:** 10.1101/2025.06.24.25330212

**Authors:** Katherine E. Brown, Jesse O. Wrenn, Nicholas J. Jackson, Michael R. Cauley, Benjamin Collins, Laurie Lovett Novak, Bradley A. Malin, Jessica S. Ancker

**Affiliations:** Department of Biomedical Informatics, Vanderbilt University Medical Center, Nashville, Tennessee; Department of Emergency Medicine, Vanderbilt University Medical Center, Nashville, Tennessee; Department of Biostatistics, Vanderbilt University Medical Center, Nashville, Tennessee; Department of Computer Science, Vanderbilt University, Nashville, Tennessee

**Keywords:** artificial intelligence, machine learning, human-AI collaboration

## Abstract

**Objective:** Healthcare decisions are increasingly made with the assistance of machine learning (ML). ML has been known to have unfairness – inconsistent outcomes across subpopulations. Clinicians interacting with these systems can perpetuate such unfairness by overreliance. Recent work exploring ML suppression – silencing predictions based on auditing the ML – shows promise in mitigating performance issues originating from overreliance. This study aims to evaluate the impact of suppression on collaboration fairness and evaluate ML uncertainty as desiderata to audit the ML.

**Materials and Methods:** We used data from the Vanderbilt University Medical Center electronic health record (n = 58,817) and the MIMIC-IV-ED dataset (n = 363,145) to predict likelihood of death or ICU transfer and likelihood of 30-day readmission. Our simulation study used gradient-boosted trees as well as an artificially high-performing oracle model. We derived clinician decisions directly from the dataset and simulated clinician acceptance of ML predictions based on previous empirical work on acceptance of CDS alerts. We measured performance as area under the receiver operating characteristic curve and algorithmic fairness using absolute averaged odds difference.

**Results:** When the ML outperforms humans, suppression outperforms the human alone (p < 0.034) and at least does not degrade fairness. When the human outperforms the ML, suppression outperforms the human (p < 5.2 × 10^-5^) but the human is fairer than suppression (p < 0.0019). Finally, incorporating uncertainty quantification into suppression approaches can improve performance.

**Conclusion:** Suppression of poor-quality ML predictions through an auditor model shows promise in improving collaborative human-AI performance and fairness.

## Introduction

Clinical decisions are increasingly made with the assistance of machine learning or artificial intelligence (ML) in the form of clinical decision support (CDS). This is often referred to as human-AI/ML collaboration (1–6). In such collaboration scenarios, the clinician is the primary decision-maker, and the AI agent processes available data and presents its prediction as supplementary information to augment existing data in the electronic health record (EHR) and prior experience of the clinician. Despite the potential for ML to equal, and possibly surpass, physicians and other clinicians in performance (7–9), many ML algorithms and systems have been known to be inconsistent in performance across subpopulations, creating what is known as algorithmic unfairness (10–12). Of further concern, recent work has shown that clinicians interacting with such systems may perpetuate algorithmic unfairness within their own decision-making by over-relying on the ML system and failing to recognize predictions with and without bias (13). Over-reliance on decision-support technology such that human performance is harmed has been studied previously in AI and non-AI tools (14,15). However, this over-reliance may become more impactful as the scope and power of ML continues to grow. This prompts the investigation of strategies to relieve clinicians from the cognitive burden of differentiating benign ML information from potentially harmful ML information.

Recent studies have begun to explore ML suppression – concealing selected ML predictions from the decision maker – as a way to mitigate overreliance (16). In this paradigm, ML decisions are not shown to the clinician if they are believed to be unreliable, incorrect, or likely to weaken the collaboration’s overall performance. If these results led to an implementation in which certain risk predictions were actually concealed from the provider, more research would be needed to understand potential impacts on provider trust, cognition, and decision making (16). Suppression could lower the likelihood the possibility that a likely faulty ML prediction is erroneously accepted by a clinician. At first glance, this solution seems to offer the best of both worlds: incorporating complementary-to-superior ML decision-support while minimizing the possibility that a human user accepts an erroneous prediction.

Although previous work by Wang et al. (17) shows that ML suppression offers some promise, this previous study 1) did not assess the effect of suppression on the overall collaboration performance and 2) did not consider the impact of ML suppression on the outcomes of patients across subpopulations. There is concern that ML suppression may inadvertently perpetuate algorithmic unfairness in collaborative performance (18). For example, it may induce a novel source of predictive unfairness by suppressing information disproportionately for specific subgroups. However, suppression could also improve fairness by suppressing erroneous predictions that disproportionately impact those subgroups. To understand these dynamics, it is important to evaluate both the performance and fairness of ML suppression.

The core challenge to ML suppression is the need to identify instances *a priori* for which the ML may be incorrect. An auditing framework may be able to support this use case (19,20). One auditing strategy is to use a secondary ML model to identify which predictions are likely to be incorrect. However, since this strategy is derived from data the ML has seen, it is reasonable to be concerned that such a strategy may not be effective for data the model has not seen in production. Alternatively, uncertainty quantification measures instability in an ML model (21,22), and when properly calculated, high uncertainty can also serve as an indicator of likely incorrectness or overall unreliability(23).

To the best of our knowledge, there have been no studies on the impact of ML suppression on the fairness of human-ML collaboration. In this work, we perform a computer-driven simulation of human-ML collaboration with ML suppression. We address the following research questions:

- RQ 1. Does selectively suppressing the AI in a collaboration scenario result in fairer or higher performing predictions than either not suppressing the ML or just relying on the human’s decisions alone?
- RQ 2. Given the performance and fairness of the human and ML, can we determine how suppression will impact performance or fairness?
- RQ 3. What benefits, if any, emerge from using uncertainty quantification of an ML model to enable suppression in human-ML collaboration?

To evaluate these questions, we built models to predict ED triage and ED discharge in data from two hospitals. We implemented gradient boosting tree (GBT) models as well as simulated oracle models that, for comparison, were made artificially more accurate. Then, we constructed ML suppression strategies that leveraged *auditor models* to identify and suppress predictions in the ML models that were likely to be erroneous or have high uncertainty. To simulate human-ML collaborative decision-making, we developed evidence-informed models that produced decisions in situations when the human decision-maker saw all ML predictions and in situations when ML recommendations with high likelihood of error or uncertainty were suppressed. Finally, we assessed the performance and fairness of these human-ML collaborations.

## Materials and Methods

This study was approved by the IRB at Vanderbilt University Medical Center (VUMC) (# 240578). We broadly consider two tasks using data from two different institutions. In the first task, *Emergency Department (ED) Triage*, the ML model predicts whether an ED patient at triage is likely to experience a negative outcome – defined as being admitted to an intensive care unit (ICU) within 24 hours or expire during the encounter. In the second task, *ED Discharge*, the ML model predicts if, at the time of ED patient discharge, the patient is likely to be readmitted to the ED within 30 days of current discharge (24,25). Tables S3 and S4 provide outcome distributions for datasets from both hospitals.

For all datasets, we follow the data preprocessing steps defined by Xie and colleagues (26) to collect patient demographics (age, sex, and race), triage vital signs, number of ED visits, hospitalizations, and ICU admissions in the past 1, 3, 7, 30, 90, and 365 days, ESI score, and 10 most frequent chief complaints. Missing demographic features were encoded as ‘Other/Unknown’, and missing triage vital signs were encoded with –1. Tables S1 and S2 provide further details about this dataset. Tables S1 and S2 provide information about the features for data from both hospitals.

## Datasets

### MIMIC-IV Data

The first two datasets are derived from the MIMIC-IV ED (27) and MIMIC-IV (28) databases (see Table S4 for cohort information). These databases are composed of data from Beth Israel Deaconess Medical Center (BIDMC) in Boston, MA. MIMIC-IV contains electronic health record (EHR) data for approximately 430,000 de-identified hospital admissions, while MIMIC-IV ED contains approximately 425,000 de-identified ED stays. Hospital admissions resulting from an ED stay can be linked to use final discharge status in predictive modeling.

For the ED Triage task, we excluded patients who either 1) were lost to care by leaving against medical advice or external transfer or 2) whose ED stays did not have an admission identifier to crosslink to a hospital stay or did not have an assigned Emergency Severity Index (ESI) score. ESI ranges from 1 indicating emergent need for medical intervention to 5 indicating a patient is stable to wait (29), and a recent study highlighted the potential for patients triaged using ESI to be mistriaged (30). The extraction process yielded *n = 393,576* patients for analysis. For the ED Discharge task, we excluded patients either 1) lost to care by leaving against medical advice or external transfer or 2) whose ED stays had an ‘Expired’ disposition. This process yielded 363,145 patients for analysis.

### VUMC Data

The other two datasets are derived from the VUMC EHR (see Table S4 for cohort information). We collected VUMC Adult ED visits between January 1, 2023, and December 31, 2023 from VUMC’s Clarity data warehouse. The total number of patients is *75,488*.

For the ED Triage task, we removed patients lost to care by leaving against medical advice or through transfer to an external facility or transfer for psychiatric evaluation, yielding 60,361 records. Approximately 94.1% of these patients did not experience a negative outcome. For the ED Discharge task, we additionally excluded patients who expired in the ED and patients in hospice, leaving 58,827 records. We filtered these patients based on their discharge dispositions from the ED and hospital. Approximately 52.3% of the patients were not readmitted to the ED within 30 days.

## Machine Learning

We relied upon CatBoost to implement GBT as our primary machine learning algorithm for these tasks (31). We performed preliminary experiments to optimize parameters for tree depth and coefficient for the L2 regularization term of the ensemble. In doing so, we applied a grid search with 3-fold cross-validation implemented within CatBoost (32). Table S5 provides the optimized values for these parameters. In our experiments, we used 1,000 maximum trees and utilized 15% of the training data as a validation set to select the number of trees that resulted in the ensemble that minimized binary log loss.

We created an artificially accurate model by performing a lookup in the dataset and returning the correct answer with a 95% likelihood. This process, often used in human-computer interaction studies, is referred to as creating an oracle model (33,34). Thus, the GBT model represents a realistic ML model that could be used in a clinical context, whereas the oracle represents a nearly best possible scenario ML performance. This allows us to consider the impact of ML performance on the overall collaboration.

## ML Suppression with Auditors

For our ML Suppression scenarios, we developed a suppression strategy that relied on **<u>auditor models</u>** to identify predictions likely to have high uncertainty or be incorrect. Table 1 lists each suppression strategy, auditor model, and identifying key used in this study.

**Table 1.** Enumeration of each auditing strategy to determine which predictions should be suppressed in the collaboration scenarios with suppression. The suppression strategy pertains to whether predicted model uncertainty or predicted model error should inform suppression. The auditor model is the underlying algorithm detecting whether an input is likely to have high uncertainty or error, respectively.

| Name | Suppression Strategy | Auditor Model |
| --- | --- | --- |
| Collaboration1 | Predict error | Logistic Regression |
| Collaboration2 | Predict error | QUEST |
| Collaboration3 | Predict uncertainty | QUEST |

We utilize a prediction of ML uncertainty, calculated using virtual ensembles (22), due to the idea that well-calibrated uncertainty is expected to be high for likely incorrect predictions and low for likely correct predictions (21,23). To train the auditor model, we use the model’s predictions on the model’s training set to develop a training set for the auditor with the outcome being whether the model was incorrect or has high uncertainty. Real-valued uncertainty is discretized into high/low categories such that each category has roughly the same number of points.

### Auditor Models

We use two types of auditor models to implement the suppression strategies. First, we use Logistic Regression (LR) with L2 penalty and regularization coefficient ! = 1 as a baseline auditor model. As input, we use the model’s predicted probability of the positive class (i.e., negative outcome/readmission within 30 days), the human’s prediction, and the ML model’s uncertainty. These inputs are used to predict the output associated with each suppression strategy. Second, we use the Quantifying Uncertainty for Estimating Subgroup Types (QUEST) ML auditing framework (23). QUEST uses pruned decision trees to estimate epistemic uncertainty; however, in addition to using model uncertainty, we also evaluate the performance of QUEST at predicting the error of the ML.

## Models of ML Collaboration

In this study, we rely upon several assumptions to model the results of the human-ML collaboration as shown in Figure 1.

**Figure 1.**
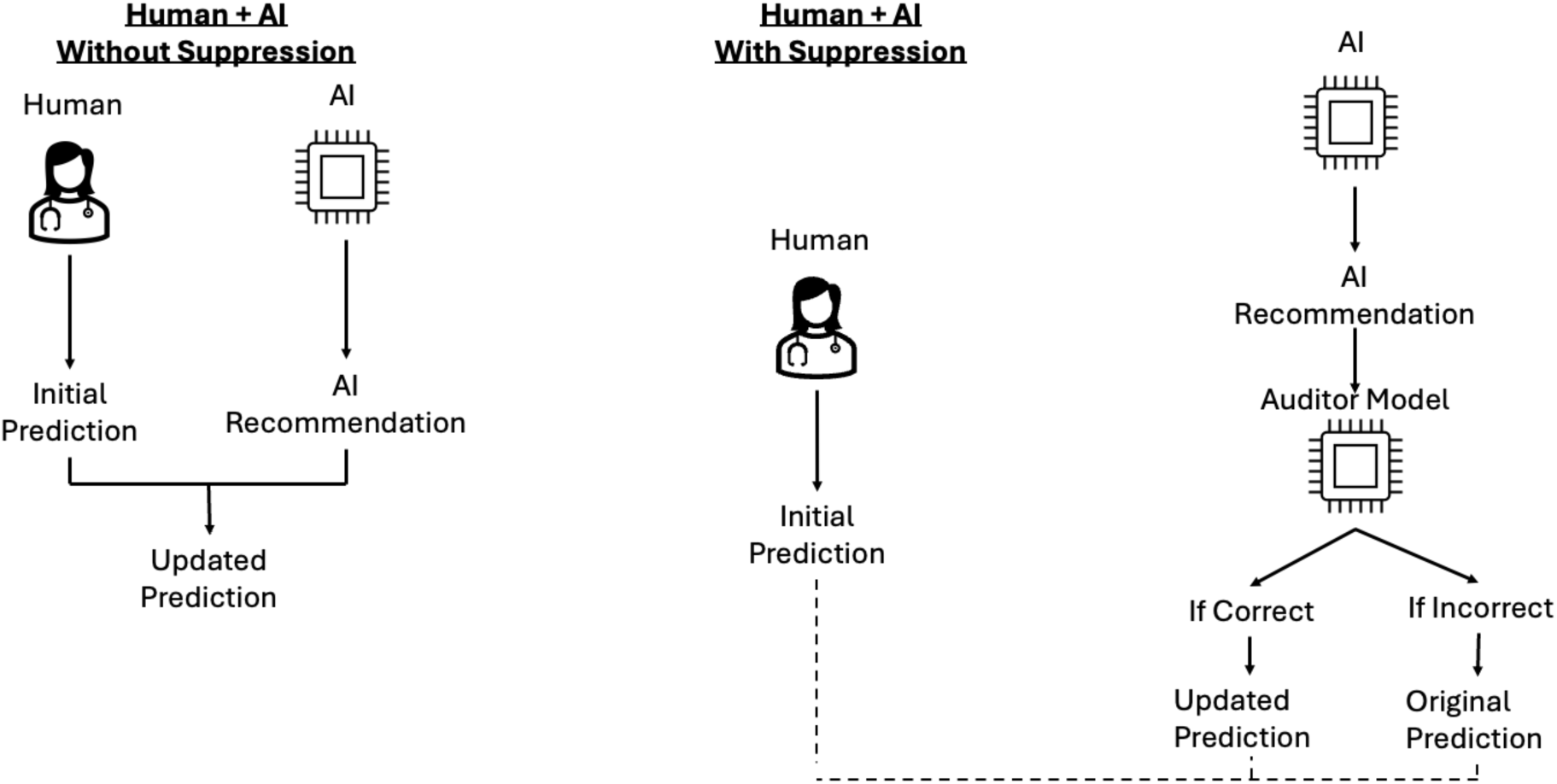
Schematic indicating the collaboration scenario with and without suppression.

For the ED Triage scenario, the documented ESI scores in the data set reflect the actual clinician triage decisions. We used our ML model and the documented ESI to predict if the patient was likely to experience a negative outcome. **Human-AI/ML Collaboration (ED Triage):** We assumed that when the model predicted a negative outcome, an alert would be presented to the clinician, and that the clinician would then update their decision in the following ways: an initial ESI of 2 would be lowered (i.e., more severe) to 1, and an initial ESI of 3, 4, or 5 would be lowered to 2.

For the ED Discharge scenario, we used our ML model at the time of patient ED discharge to produce a prediction of likelihood of 30-day readmission (Figure 1). Our model did not incorporate the actual discharge disposition.

**Human-AI/ML Collaboration (ED Discharge):** We assumed that when the model predicted a high likelihood of readmission, the clinician would change their discharge evaluation and admit the patient instead of discharging them.

**Human-AI/ML Collaboration Model modification – CDS overrides:** For both models, we made an additional set of adjustments on the basis of empirical work on how human decision makers respond to CDS. Previous work by Sittig et al. (35) suggests that clinicians are more likely to accept alerts on elderly patients (> 65 years), patients currently on at least five medications, and those with five or more chronic conditions. In recognition of the reality that CDS alerts are not always utilized (36), we intentionally reduced the acceptance rate of the alerts for the ML models. Based on input from our clinical domain expert (JW), we also considered the impact of the severity of the patient’s primary complaint. Thus, if the patient is elderly, polypharmacy (requires multiple medications), has five or more chronic conditions, or fulfills the severity assumption of their chief complaint, we modeled the likelihood of accepting the CDS suggestion as 100%. If a patient does not meet any of these criteria, we modeled a 20% acceptance rate for the CDS based on the output of a numPy random number generator library in Python (37).

For the remainder of this paper, human-AI/ML collaboration without suppression will be denoted as CollaborationAll, where the ML-AI is shown for all predictions. Further, human-AI/ML collaboration with suppression will be referred to as Collaboration1, Collaboration2, or Collaboration3 (see Table 1).

## Performance and Fairness Evaluation

To evaluate the performance of the simulated collaborations, we report both the area under the receiver operator characteristic (AUROC) curve and area under the precision-recall curve (AUPRC). When applicable, in the ED Triage task, we derive these curves based on thresholds derived from the ESI scores.

To evaluate the fairness of the simulated collaborations, we use absolute average odds difference (AAOD) as our primary fairness metric. AAOD is a measure based on the equalized odds (EO) measure of algorithmic fairness (38). In EO, a classifier is considered fair when, given a majority subgroup *s_maj_* and a minority subgroup *s_min_*, the following is true: 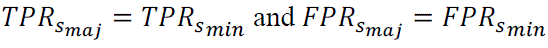, where TPR represents true positive rate and FPR represents false positive rate. AAOD combines these conditions of EO into one distinct formula by averaging differences in TPRs and FPRs across the majority and minority subgroups. Specifically, AAOD is defined as 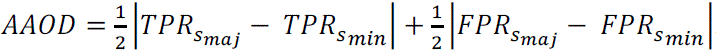. For both datasets, we consider subgroups (majority/minority) with respect to race (white/not white), age (less than 50 years old/50 years old or older), and gender (male/not male).

## Experimental Design and Statistical Analysis

We evaluate our simulated collaboration scenario by partitioning the target dataset into a 75%/25% training set/testing set. We train our ML model and the auditor model on the training set and evaluate the ML and auditor models on the testing set. We report the performance on the testing set. We repeat this entire procedure (including data partitioning and model training) a total of 30 times to have enough samples for statistical testing. We report average and 95% confidence intervals for the performance of the ML model, human, and simulated collaborations. We also report AAOD for the ML model, human, and collaborations. We use the sum of the AAOD values for each of the demographic columns and compute mean and 95% confidence intervals. To determine statistical significance, we apply the Mann-Whitney U-test due to the observation that some of the AUROC and AAOD distributions are skewed and/or bimodal. Our null and alternative hypotheses for the statistical tests are as follows: Let 1_’_ and 1_(_denote two models (i.e., ML predictions, human predictions, collaboration without suppression, or collaboration with suppression (see Table 1). For performance, our null hypothesis is that there is no difference in central tendency between the distribution of AUROC values of the models, and our alternative hypothesis is that the median of AUROC of 1_’_ is greater than that of 1_(_. Similarly, for fairness, our null hypothesis is that there is no difference in central tendency between the distribution of AAOD values of the models, and our alternative hypothesis is that the median of AUROC of 1_’_ is less than that of 1_(_.

## Results

Table 2 reports on the AUROC and AUPRC for each model. Figures 2 and 3 evaluate the fairness-performance trade-off, with performance on the X-axis and fairness on the Y-axis. Finally, we present the p-values of the Mann-Whitney U tests in Figures 4 and 5.

**Figure 2.**
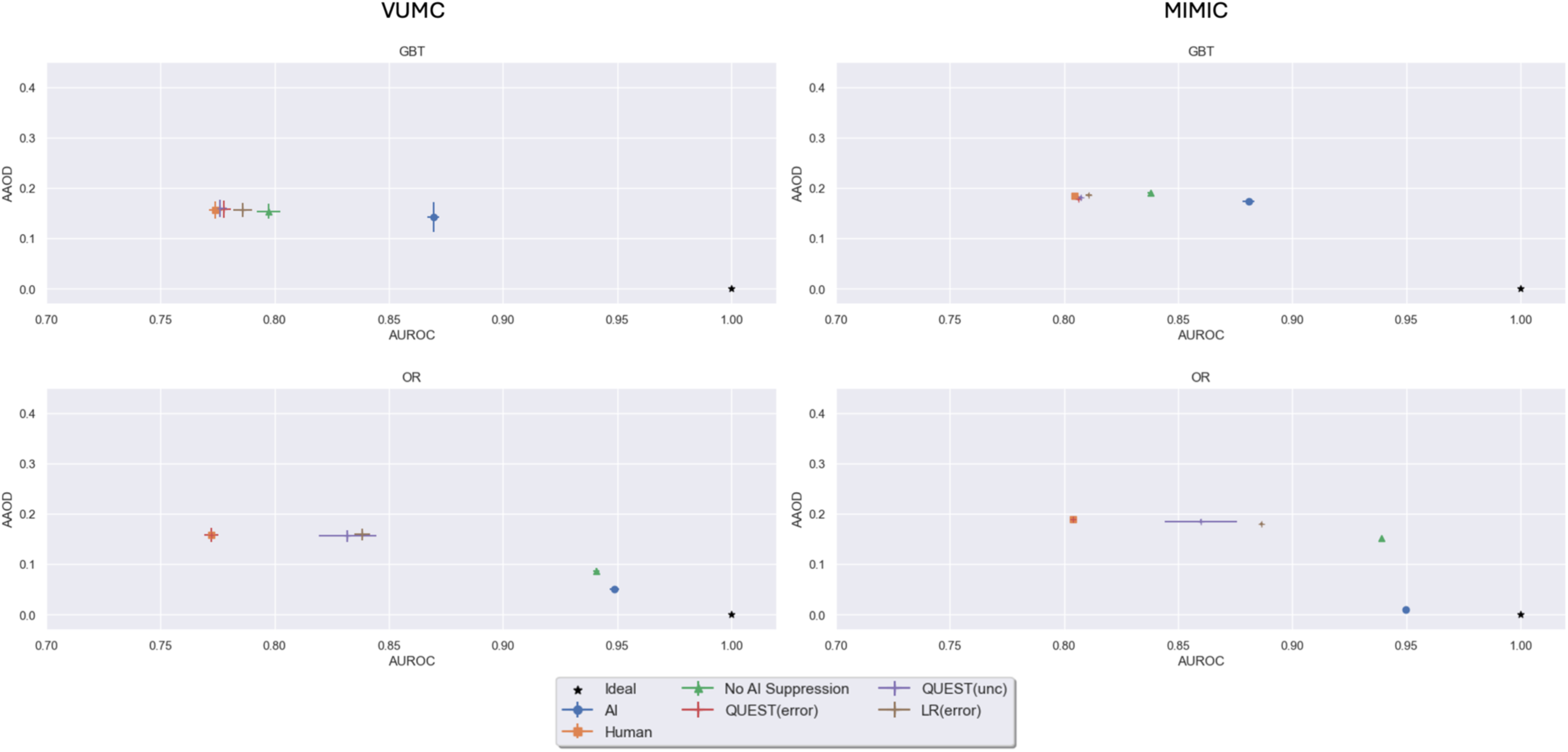
Fairness-utility tradeoff plots depicting the average absolute odds difference on the y-axis and the performance in area under the ROC curve on the x-axis. Error bars depicting 95% CI are included. Prediction task: ED Triage.

**Figure 3.**
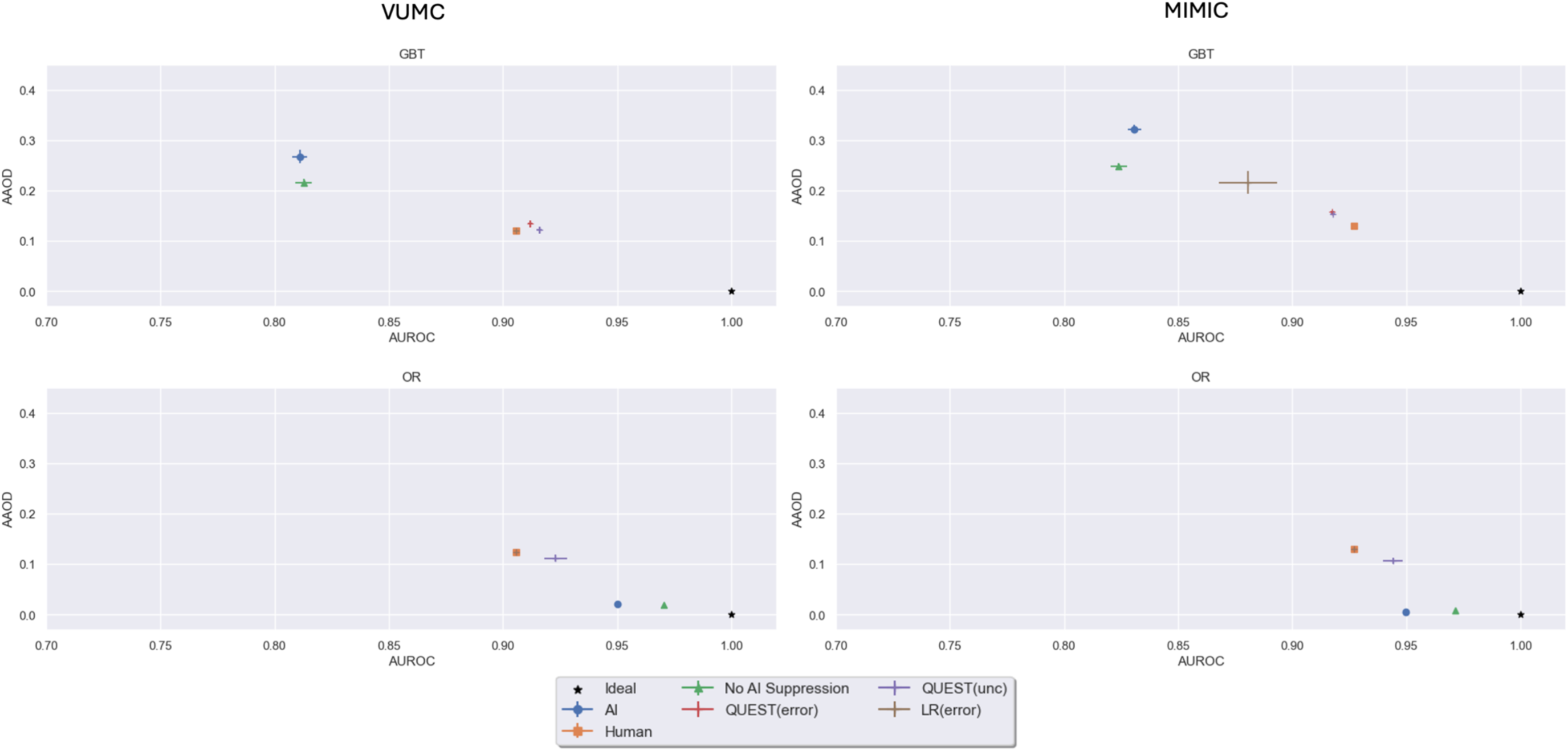
Fairness-utility tradeoff plots depicting the average absolute odds difference on the y-axis and the performance in area under the ROC curve on the x-axis. Error bars depicting 95% CI are included. Prediction Task: ED Discharge.

**Figure 4.**
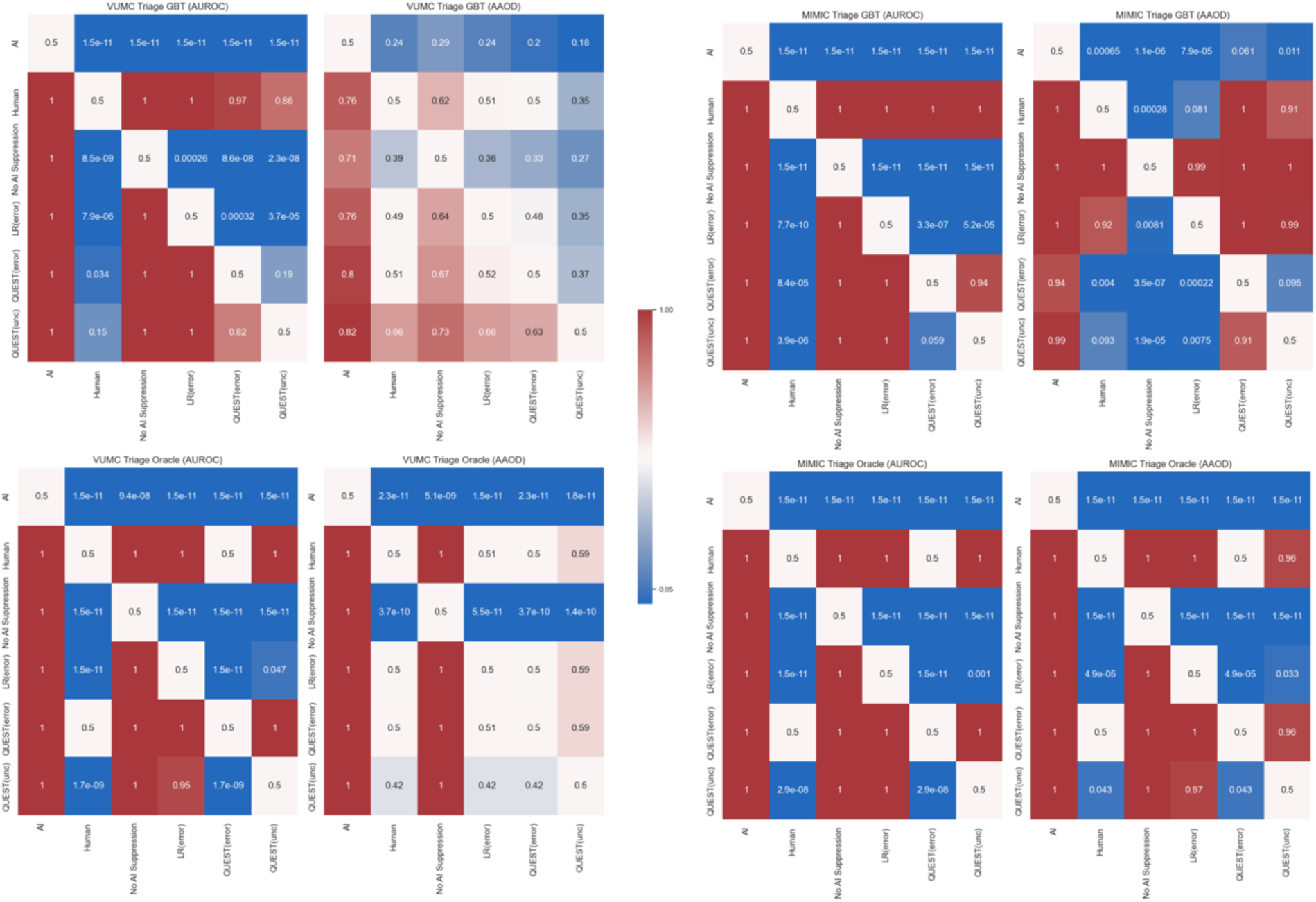
Heatmap of p-values resulting from the Mann-Whitney U Test for statistical significance. The p-value is for the test that the model given by the row is higher performing or fairer than the model given by the column. Task: ED Triage.

**Figure 5.**
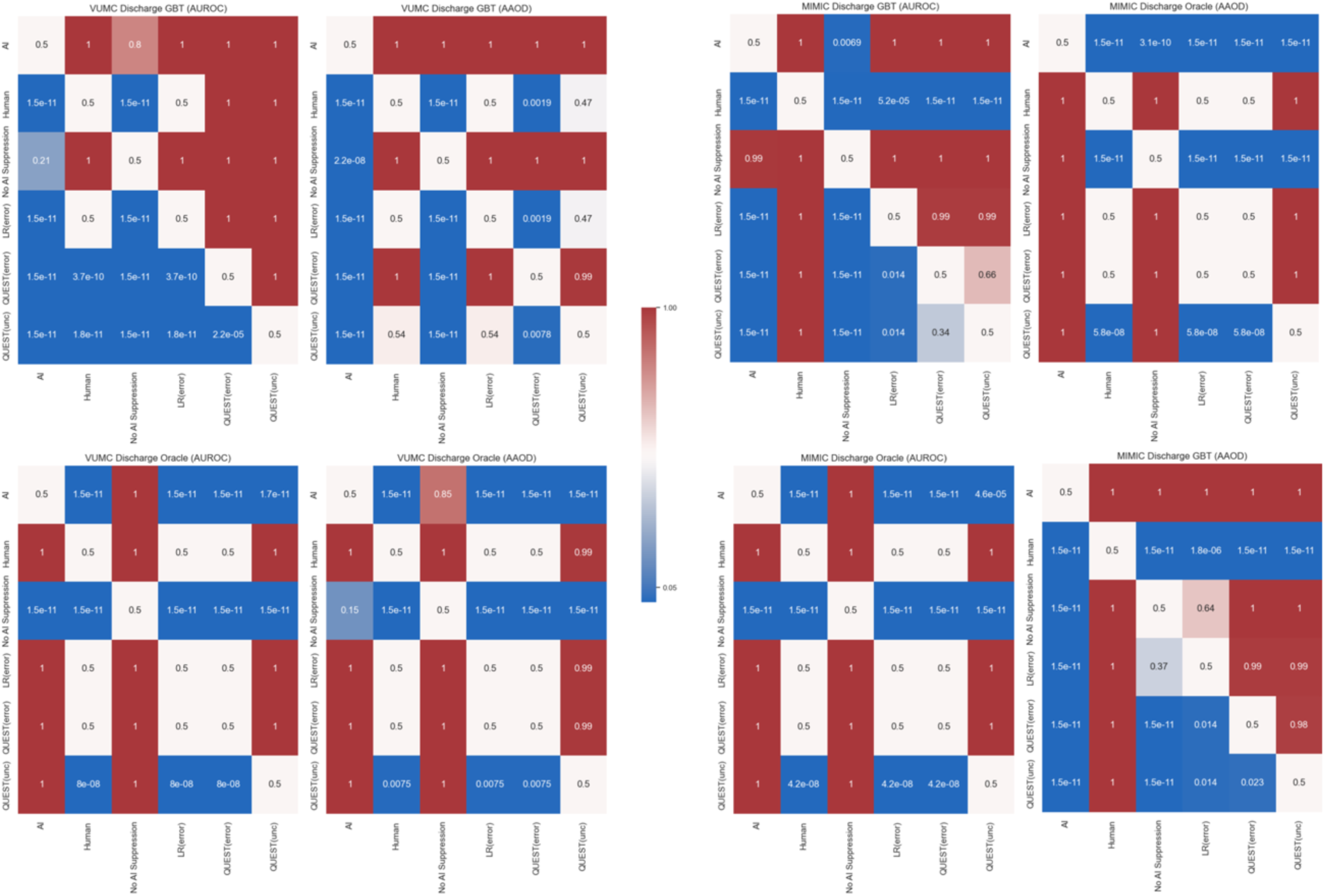
Heatmap of p-values resulting from the Mann-Whitney U Test for statistical significance. The p-value is for the test that the model given by the row is higher performing or fairer than the model given by the column. Task: ED Discharge

**Table 2.** Mean (95% CI) of the area under the receiver-operating characteristic (ROC) and precision-recall (PRC) curves. We evaluate each task, model or simulated collaboration scenario and differentiate between a GBT AI and the oracle AI.

|  |  | GBT |  | Oracle |  |
| --- | --- | --- | --- | --- | --- |
|  | Model | AUROC | AUPRC | AUROC | AUPRC |
| VUMC<br>Triage | AI | 0.87 (0.867, 0.872) | 0.324 (0.314, 0.333) | 0.949 (0.947, 0.951) | 0.441 (0.435, 0.448) |
|  | Human | 0.774 (0.771, 0.777) | 0.185 (0.181, 0.189) | 0.772 (0.769, 0.776) | 0.183 (0.179, 0.186) |
|  | Collaboration w/o Suppression | 0.797 (0.792, 0.802) | 0.186 (0.182, 0.191) | 0.941 (0.939, 0.942) | 0.321 (0.317, 0.324) |
|  | Simple(error)-LR | 0.786 (0.782, 0.79) | 0.186 (0.181, 0.19) | 0.838 (0.835, 0.842) | 0.267 (0.263, 0.272) |
|  | Simple(error)-QUEST | 0.778 (0.775, 0.781) | 0.188 (0.184, 0.192) | 0.772 (0.769, 0.776) | 0.183 (0.179, 0.186) |
|  | Simple(unc)-QUEST | 0.776 (0.773, 0.779) | 0.189 (0.184, 0.193) | 0.832 (0.819, 0.845) | 0.255 (0.241, 0.269) |
| VUMC<br>Readmit | AI | 0.906 (0.905, 0.906) | 0.802 (0.798, 0.805) | 0.95 (0.95, 0.951) | 0.932 (0.931, 0.933) |
|  | Human | 0.906 (0.905, 0.906) | 0.951 (0.95, 0.951) | 0.906 (0.905, 0.907) | 0.951 (0.95, 0.951) |
|  | Collaboration w/o Suppression | 0.813 (0.809, 0.816) | 0.849 (0.847, 0.851) | 0.971 (0.97, 0.971) | 0.972 (0.971, 0.972) |
|  | Simple(error)-LR | 0.906 (0.905, 0.906) | 0.951 (0.95, 0.951) | 0.906 (0.905, 0.907) | 0.951 (0.95, 0.951) |
|  | Simple(error)-QUEST | 0.912 (0.911, 0.913) | 0.941 (0.94, 0.942) | 0.906 (0.905, 0.907) | 0.951 (0.95, 0.951) |
|  | Simple(unc)-QUEST | 0.916 (0.915, 0.917) | 0.947 (0.945, 0.948) | 0.923 (0.918, 0.928) | 0.958 (0.956, 0.96) |
| MIMIC<br>Triage | AI | 0.881 (0.878, 0.883) | 0.414 (0.405, 0.422) | 0.95 (0.949, 0.95) | 0.551 (0.549, 0.553) |
|  | Human | 0.805 (0.804, 0.805) | 0.38 (0.379, 0.382) | 0.804 (0.803, 0.804) | 0.379 (0.377, 0.381) |
|  | Collaboration w/o Suppression | 0.838 (0.837, 0.839) | 0.471 (0.469, 0.474) | 0.939 (0.939, 0.939) | 0.688 (0.687, 0.689) |
|  | Simple(error)-LR | 0.811 (0.81, 0.812) | 0.394 (0.39, 0.398) | 0.886 (0.886, 0.887) | 0.592 (0.59, 0.593) |
|  | Simple(error)-QUEST | 0.806 (0.806, 0.807) | 0.383 (0.381, 0.384) | 0.804 (0.803, 0.804) | 0.379 (0.377, 0.381) |
|  | Simple(unc)-QUEST | 0.807 (0.807, 0.808) | 0.381 (0.38, 0.383) | 0.86 (0.844, 0.876) | 0.513 (0.476, 0.549) |
| MIMIC<br>Readmit | AI | 0.831 (0.828, 0.834) | 0.847 (0.845, 0.85) | 0.95 (0.95, 0.95) | 0.939 (0.938, 0.939) |
|  | Human | 0.927 (0.927, 0.927) | 0.964 (0.964, 0.964) | 0.927 (0.927, 0.927) | 0.964 (0.964, 0.964) |
|  | Collaboration w/o Suppression | 0.824 (0.82, 0.828) | 0.87 (0.868, 0.872) | 0.971 (0.971, 0.972) | 0.975 (0.975, 0.975) |
|  | Simple(error)-LR | 0.881 (0.868, 0.893) | 0.916 (0.903, 0.929) | 0.927 (0.927, 0.927) | 0.964 (0.964, 0.964) |
|  | Simple(error)-QUEST | 0.918 (0.917, 0.919) | 0.949 (0.948, 0.95) | 0.927 (0.927, 0.927) | 0.964 (0.964, 0.964) |
|  | Simple(unc)-QUEST | 0.918 (0.917, 0.919) | 0.949 (0.948, 0.95) | 0.944 (0.94, 0.949) | 0.969 (0.968, 0.97) |

## Performance and Fairness of Human and ML Separately

For the ED Triage task, the GBT achieved statistically significantly higher performance in AUROC (p < 1.5 × 10^-11^) and better fairness according to AAOD (p < 0.0065) than the documented human ESI values. However, the difference was not statistically significant for fairness in the VUMC data (p > 0.18). The oracle statistically significantly outperforms (p < 1.5 × 10^-11^) and is fairer (p < 2.3 × 10^-11^) than the human for both datasets. For the ED Readmission task, in both datasets, the human statistically significantly outperformed the GBT with respect to performance (VUMC: p < 1.5 × 10^-11^, MIMIC: p< 1.5 × 10^-11^) and fairness (VUMC: p < 1.5 × 10^-11^, MIMIC: p< 1.5 × 10^-11^). The oracle statistically significantly outperformed the human with respect to performance (VUMC: p < 1.5 × 10^-11^, MIMIC: p< 1.5 × 10^-11^) and fairness (VUMC: p < 1.5 × 10^-11^, MIMIC: p < 1.5 × 10^-11^).

## Collaboration Performance and Fairness without Suppression

For the ED Triage task, in both datasets, the GBT outperforms CollaborationAll (VUMC: p < 1.5 × 10^-11^, MIMIC: p < 1.5 × 10^-11^) collaboration. However, the GBT is fairer than the CollaborationAll in only the MIMIC dataset (p < 1.1 × 10^-6^). By contrast, for the ED Readmission task with the oracle, the Collaboration All outperforms the oracle alone for both datasets (VUMC: p < 1.5 × 10^-11^, MIMIC: p < 1.5 × 10^-11^). This is likely due to the lower performance of the human in the Triage task as opposed to the Readmission task. We suspect that the patients frequently misclassified by the human are those for whom the rules for human decision override do not apply.

## Impact of Suppression on Collaboration Performance and Fairness

There is one scenario, namely when GBT is applied to the VUMC Readmit task, in which incorporating ML suppression (i.e., CollaborationX) has higher AUROC than the other predictors. Moreover, this finding is statistically significant (p < 1.5 × 10^-11^). In this scenario; however, CollaborationX with GBT only improved upon the AI’s fairness (p <1.5 × 10^-11^) and not the human’s fairness. Using Collaboration3 produced the highest performance of all suppression techniques (p < 2.2 × 10^-5^), but this technique (p < 0.0078) and the Collaboration1 (p < 0.0019) were both statistically significantly fairer than Collaboration2.

Since ML suppression does not uniformly improve AUROC or fairness, we compared collaboration with ML suppression to the human. We find that Collab-AI-Suppressed statistically significantly improves upon the human’s AUROC for all evaluated dataset and model combination evaluated (p < 0.034) except when GBT is applied to the MIMIC Readmit task. The human statistically significantly outperforms the CollaborationX for this task (p < 5.2e-5). Additionally, we also observed that when the ML has higher AUROC than the human, a form of ML suppression (CollaborationX) outperformed the human (p < 0.034) with either no detriment to fairness or improvement in fairness (p < 0.075). However, if the human has higher AUROC than the ML, we notice that the human is statistically significantly fairer than all forms of ML suppression (CollaborationX, p < 0.0019). Thus, we believe that ML suppression has the potential to improve performance over the human alone in the right circumstances.

Finally, we compare the ML suppression techniques to determine if there are any advantages to one technique over the others (Table 3). Collaboration1 was most frequently the highest performing suppression technique; whereas, Collaboration3 was frequently the AI suppression technique that resulted in the fairest collaboration.

**Table 3.** Counts of the number of time each suppression strategy was the highest performing or fairest among all 3 suppression strategies. The number of times a strategy is statistically significantly best is given in parenthesis.

|  | Performance | Fairness |
| --- | --- | --- |
| LR(error) | 4 (4) | 2 (1) |
| QUEST(error) | 0 | 1 (0) |
| QUEST(unc.) | 3 (3) | 4 (3) |
| Tie | 1 | 1 |

In summary, these findings show that when the human has higher AUROC than the ML, collaboration with ML suppression outperforms collaboration without ML suppression, and when the ML is higher performing, collaboration without ML suppression outperforms collaboration with AI suppression.

## Discussion

This simulation study ML suppression strategies and human-ML collaborations indicated that collaboration with ML suppression can improve performance over humans alone. In many, although not all, instances this improvement comes with an improvement in fairness (RQ 1). We observed that when the ML is higher performing compared to the human, a form of suppression will be either as fair or fairer than the human. Conversely, when the human is higher performing than the ML, the human will be fairer than any form of suppression (RQ 2).

Additionally, when we compare collaboration with suppression to collaboration without suppression, we find for a realistic AI (i.e., GBT), collaboration with suppression is often fairer than collaboration without suppression (RQ 1). Similarly, we find suppression benefits the collaboration more than not having suppression when the human is higher performing, and no suppression is preferable to suppression when the ML is higher performing (RQ 2). Finally, our experiments suggest that uncertainty quantification could be a useful tool to benefit human-ML collaboration (RQ 3).

Despite its potential, one concern with ML suppression is that poor implementation could unjustly disadvantage certain subpopulations (18). For instance, such an implementation could learn that an ML has lower performance for a specific subpopulation and ultimately suppress predictions for that subpopulation. This would result in some subpopulations receiving the benefits of ML while others do not. We considered an AI implemented in good faith (GBT) and an simulated ML with near perfect performance (oracle model) and, in our experiments, these concerns were not realized. For the GBT– which represents a realistic ML – the ML suppression was fairer in most circumstances. We believe that future work should compare the impact of AI suppression on collaborations with higher-performing, lower-performing, and systematically biased ML.

Uncertainty quantification has currently been under-studied with respect to human-ML collaboration, and we posit that leveraging ML uncertainty in suppression could be an effective tool to assist with human-AI collaboration. We further noticed that using uncertainty quantification as either 1) input into the auditor or 2) as the output of the auditor to drive the suppression decision resulted in improved performance and fairness, respectively. As an illustration, we noticed that using uncertainty to inform the decision of whether to suppress the ML resulted in the fairest suppression approach in four out of eight experiments. This is well-aligned with previous studies that suggest AI uncertainty can be used to help distinguish between subpopulations for which an ML model has higher and lower fairness (39), as well as performance (23).

Another aspect of ML suppression that we did not consider is how users interpret the presence – or lack thereof – of a CDS alert. Evidence suggests that people can change their mind based on AI or CDS responses and behavior (35,40,41). A prediction that is displayed only part of the time may be ignored by the users. On the other hand, if users know, or learn over time, that the lack of an ML –based alert is associated with the ML being incorrect, then this could reinforce existing automation bias.. As a consequence, the ML could drift – degrade in either performance or fairness – in the future due to changes in patient populations or other clinical changes. If the auditor is not routinely updated, it could fail to suppress incorrect predictions resulting from a drift in performance, and as a result, the automation bias resulting from assuming an ML alert is correct could induce a degradation in performance.

Thus, it is important for the research and application communities to understand how users interact with auditor-based systems and how auditor models respond to model drift.

There are several limitations of our study we wish to acknowledge. First, we simulated human decision-making rather than evaluating actual human responses from clinicians. Future work is needed to elucidate the impacts of *how* suppression is implemented within the user interface and the impact on clinician trust and reliance on the system. Additionally, our analysis uses retrospective data. The scenario could exist, for example, where a patient received low (i.e., more severe) triage score and received rapid intervention that prevented a negative outcome, causing a mismatch between outcome and triage score. To mitigate this for this work, we only allowed predictions to increase in severity; however, to fully account for this, a prospective study would be required.

## Conclusion

Suppression of poor-quality ML predictions through an auditor model shows promise in improving collaborative human-AI performance and fairness. When the ML outperforms humans, suppression improves performance and improves – or at least does not degrade – fairness. When the human outperforms the ML, suppression improves performance but does not improve collaboration fairness. Finally, incorporating uncertainty quantification into suppression approaches can in some cases improve performance. Future research should verify these findings without simulation on additional retrospective and prospective experiments.

## DATA AVAILABILITY

The MIMIC-IV dataset underlying this article are available in Physionet at https://doi.org/10.13026/5ntk-km72. The VUMC data underlying this article cannot be shared publicly to preserve the privacy of individuals in the study. The data are available from the corresponding authors on reasonable request.

## AUTHOR CONTRIBUTIONS

Using the CRediT Taxonomy, the following are the roles of the authors on the manuscript:

KB: Conceptualization, Data Curation, Formal analysis, Investigation, Writing (original draft, review & editing)

JW: Data curation, Writing (original draft, review & editing)

NJ: Writing (review & editing) MC: Writing (review & editing)

BC: Writing (review & editing) LN: Writing (review & editing)

BM: Conceptualization, Resources, Writing (review & editing)

JA: Conceptualization, Resources, Supervision, Writing (original draft, review & editing)

