## Supplementary material for "Auditor Models to Suppress Poor AI Predictions Can Improve Human-AI Collaborative Performance": S1

S.1 Additional Dataset and Model Information

Table S1. Enumeration of features in the MIMIC-IV and MIMIC-IV ED datasets originating from Beth Israel Deaconess Medical Center. For values in between angled brackets (<…>), there is a single feature per value. Feature name, type, description, and usage per task is included.

| Feature Name | Type | Description | Clinical Task Used |
|---|---|---|---|
| No. Days in the ED in the past <1/3/730/90/365> days | Integer | Number of days patient has visited the ED in the last <1/3/7/30/90/365> | ED Triage<br>ED Discharge |
| No. Days as an Inpatient in the Hospital in the past <1/3/730/90/365> days | Integer | Number of days patient has been admitted to the hospital in the last <1/3/7/30/90/365> | ED Triage<br>ED Discharge |
| No. Days in the ICU in the past <1/3/730/90/365> days | Integer | Number of days patient has been admitted to the ICU in the last <1/3/7/30/90/365> | ED Triage<br>ED Discharge |
| Age | Float | Age of patient | ED Triage<br>ED Discharge |
| Gender | Categorical | Male/Female | ED Triage<br>ED Discharge |
| Race | Categorical | Race of patient | ED Triage<br>ED Discharge |
| Arrival Transport | Categorical | Means of arrival for patient | ED Triage<br>ED Discharge |
| Triage <Temperature/Heart Rate/Respiratory Rate/O2 Saturation/Systolic BP/Diastolic BP/Pain Level> | Continuous | Specified vital sign at triage | ED Triage<br>ED Discharge |
| Average <Temperature/Heart Rate/Respiratory Rate/O2 Saturation/Systolic BP/Diastolic BP/Pain Level> | Continuous | Average of specified vital sign during ED visit | ED Discharge |
| Last <Temperature/Heart Rate/Respiratory Rate/O2 Saturation/Systolic BP/Diastolic BP/Pain Level> | Continuous | Last measurement of specified vital sign during ED visit | ED Discharge |
| Emergency Severity Index | Categorical | 1 (most severe), 2, 3, 4, 5 (least severe) | ED Triage<br>ED Discharge |
| Chief Complaint Present | Binary | Is the specified chief complaint present in this patient? (0: No/1: Yes) | ED Triage<br>ED Discharge |
| Charleston Comorbidity Index | Binary | Is the specified comorbidity present? (0: No/1: Yes) | ED Triage<br>ED Discharge |
| Elixhauser Comorbidity Index | Binary | Is the specified comorbidity present? (0: No/1: Yes) | ED Triage<br>ED Discharge |
