## Supplementary material for "Auditor Models to Suppress Poor AI Predictions Can Improve Human-AI Collaborative Performance": S3

Table S3. Distribution of outcomes and majority and minority classes of demographics in the MIMIC-IV data across both tasks.

| | | ED Triage | | ED Discharge | |
|---|---|---|---|---|---|
| | Ground Truth | No Negative Outcome | Negative Outcome | Not Readmitted within 30 days | Readmitted within 30 days |
| Age | 18-49 | 163,574 | 4,559 | 104,275 | 51,203 |
| | >= 50 | 204,569 | 20,874 | 73,862 | 133,805 |
| Race | White | 197,421 | 15,635 | 85,619 | 110,709 |
| | Not White | 170,722 | 9,798 | 92,518 | 74,299 |
| Gender | Male | 165,434 | 13,652 | 74,251 | 89,716 |
| | Not Male | 202,709 | 11,781 | 103,886 | 95,292 |
| | Total | 368,143 | 25,433 | 178,137 | 185,008 |
