## Supplementary material for "Auditor Models to Suppress Poor AI Predictions Can Improve Human-AI Collaborative Performance": S4

Table S4. Distribution of outcomes and majority and minority classes of demographics in the VUMC data across both tasks.

| | | ED Triage | | ED Discharge | |
|---|---|---|---|---|---|
| | Ground Truth | No Negative Outcome | Negative Outcome | Not Readmitted within 30 days | Readmitted within 30 days |
| Age | 18-49 | 32,389 | 1,011 | 20,835 | 14,733 |
| | >= 50 | 23,961 | 1,456 | 9,958 | 13,297 |
| Race | White | 35,745 | 1,785 | 18,564 | 18,968 |
| | Not White | 20,605 | 682 | 12,229 | 9,066 |
| Gender | Male | 26,511 | 1,452 | 13,230 | 14,733 |
| | Not Male | 29,839 | 1,015 | 17,563 | 13,301 |
| | Total | 56,350 | 2,467 | 30,793 | 28,034 |
